# The impact of a fairytale-like story on the food choices of preschool children

**DOI:** 10.1101/2024.10.14.24314631

**Authors:** Anoushiravan Zahedi, Stephen Katembu, Sharon Michelle Sind, Undine Sommer, Charles Kimamo, Werner Sommer

## Abstract

The ongoing obesity epidemic is an indicator that traditional efforts towards diet change are insufficient, and interventions focusing mainly on restrictions of unhealthy food are of limited success. Therefore, approaches targeting food preferences should be integral in counteracting the current epidemic. However, food preferences are affected by a multitude of factors and are usually resistant to changes later in life. Hence, we tested whether the food choices of preschool children can be changed by a child-appropriate, interactive, fairytale-like narrative. We used two interactive stories: the first (experimental) story was about how two protagonists of similar age as the participants saved their hometown from being drained of color with the help of magic fruits or vegetables, while the second (control) story consisted of a similar plot that did not mention food. In Experiment 1, we used a crossover design with a one-week baseline measurement. After the experimental stories, healthy food choices (i.e., pieces of fruit vs. cookies, sweets, or cake) increased significantly relative to the one-week baseline, whereas no change was observed after the control story. In preregistered Experiment 2, we replicated these results with vegetables as healthy options using a random control design and investigated the longevity of the effects. The food-story effect on vegetable choices was similar to fruits (Exp. 1) and lasted for two weeks. These findings demonstrate that appropriate narratives about food can have a strong and lasting effect on the healthy food choices of preschool children and might promote healthy food consumption if incorporated into nutrition education.

## 1. Introduction

Food is nourishing and often pleasant. However, what and how much we eat (food choice) can become a problem for both human health (GBDRF Collaborators, 2020) and planet health (Clark et al., 2019; Willett et al., 2019). A major health concern related to food is obesity (Tiwari & Balasundaram, 2023), which is linked to many noncommunicable diseases (NDC-RisC, 2024; Weihrauch-Blüher & Wiegand, 2018). This problem is not restricted to high-income countries but is increasingly observed also in low- and mid-income countries (Lin et al., 2020). Especially troubling is the rising incidence of obesity in children and adolescents (NDC-RisC, 2024; Ogden et al., 2007). As compared to their normal-weight age-mates, obese children have a five-fold higher risk of also being obese in adulthood (Kelsey et al., 2014; Simmonds et al., 2016), and weight increase only accelerates in children and adolescents as they become older (Baum II, Ruhm, 2009). These findings indicate the necessity of early interventions and preemptive measures. Thus, due to the complexity involved in reversing weight gains as we age (Madigan et al., 2022), interventions should already target young cohorts’ food preferences and choice habits to prevent obesity by creating lifelong healthy habits (Kelsey et al., 2014). However, previous treatment attempts for childhood obesity have not shown long-term success (Al-Khudairy et al., 2017; Mead et al., 2017), showing the importance of using novel and innovative techniques. Hence, in the present study, we targeted eating behavior and, more specifically, food choice in preschool children aged 4 to 6 by using fairytale-like narratives.

Food preferences are influenced by a complex blend of genetic, environmental, and social factors (Scaglioni et al., 2011), such as the social or situational context, cultural background, the preparation of the food (Renner et al., 2012; Wahl et al., 2020), and family settings (Larsen et al., 2015). Like many affective responses and semantic world knowledge, food preferences are, to a substantial degree, culturally transmitted (Henrich, 2016). Many individuals struggle to alter their dietary habits and change their food preferences, which especially affects eating behavior when making impulsive, non-deliberate decisions (Bartkiene et al., 2019). Notably, the family has an integral role, especially in young children’s food preferences and intake habits (Rahill et al., 2020). The influence of parents on children’s food intake is multifaceted (Larsen et al., 2015) since not only food parenting behavior influences child weight and eating self-regulation (Grammer et al., 2022) but also the parent’s dietary habits may be transferred to children (Larsen et al., 2015; Rahill et al., 2020). Parental effects are, however, a double-edged sword, as not only may parents transfer unhealthy habits to their children, but they might also use ineffective parenting behavior, swhich might result in unhealthy food preferences in the children (Russell et al., 2015). Therefore, interventions that focus on [pre-]school settings are vital to creating healthy eating habits in children.

Narratives have been a creative, communicative medium that is entertaining and transmits information and cultural beliefs. According to Grainger (1997), storytelling endeavors to elucidate the mysteries of life and the world, helping to make sense of complex concepts, with characters and themes within these narratives being a means of cultural continuity. Stories reflect and shape the way we think and captivate our imagination. Narratives, ‘the natural product of language’ (Donald, 1991, p.275), are a powerful influence in early childhood and adolescent development, enhancing language acquisition, social skills, critical thinking, and moral reasoning (Isbell et al., 2004) while promoting an understanding of both community and self (Dunne, 2006). Narratives have already been used to successfully affect children’s social behavior and individual choices (Teglasi & Rothman, 2001; see Montgomery et al., 2015 for meta-analysis). Further, minimally counterintuitive narratives, such as fairytales, have been discussed as a powerful tool for the perception and reflection of emotions and choices in children (Hohr, 2000). They can facilitate emotional and cognitive development in children by promoting resilience, empathy, and socialization by encouraging children to reflect on moral choices and the consequences of actions. (Koutsompou, 2016). Therefore, as we aim to alter food choices in preschool children, fairytale-like narratives are a promising tool, which are known to be loved by preschool children and critically influence their development (Muindi, 2017).

Traditionally, narratives appear to be especially attention-catching if they contain concepts that partially violate everyday real-world knowledge, such as animals that speak or understand human language, inanimate objects that move on command, ghosts that can pass through walls, etc. (Boyer, 1994). Previous findings have shown that stories that contain a small number of counterintuitive ideas, that is, ideas that disagree to some extent with the world knowledge of the receiver (e.g., the speaking frog in the fairytale of the Frog King, Grimm & Grimm, 1812-1857) are more memorable than stories without such violations (Aristei et al., 2022; Knoop et al., 2024; Norenzayan et al., 2006). It is further discussed that the mnemonic resilience of narratives with minimally counterintuitive concepts results in their cultural success (Norenzayan et al., 2006). As Warner (2018) in ‘Fairy Tale: A Very Short Introduction’ discusses what makes a narrative an artistic fairytale is containments of minimally counterintuitive elements. Notably, in contrast to folkloric fairytales which are closely related to folk tales, artistic fairytales are usually culturally neutral, meaning their magical elements should be perceived as minimally violating core knowledge regardless of listeners’ cultural background. We use fairytale-like narratives throughout the text to refer to this concept of culturally neutral artistic fairytales.

With narratives possessing such power, the role of food narratives - stories that associate foods with characters, adventures, or moral lessons - in influencing children’s food choices appears to be especially promising. Narratives applied to food education may create positive associations with healthy foods, making them more appealing to young children. A pioneer in using stories to influence food preferences is Karl Duncker (1938), who used a story about a mouse named “Eaglefeather,” who was looking in the forest for food, to manipulate children’s food preferences. Eaglefeather came across a delicious-tasting bark called “Maple”; later, it found another bark called “Hemlock,” which it found to be distasteful. After the narrative and some playful elements, the experimenter let the children taste samples of both “foods” and asked for their preference; the “delicious Maple” was represented by sugar cubes with some drops of valerian (tasting distinctly medicinal), and the “distasteful Hemlock” was represented by white chocolate (with a nice lemon taste). In line with the narrative about Eaglefeather, the children stated a strong preference for maple over hemlock. In contrast, in a control group of children who had not heard the story, there was a clear taste preference for hemlock. The effect persisted for almost one week of repeated ratings. Although promising, there seems to be no follow-up of Duncker’s (1938) study to check the generalizability of these effects to real food or to better control for confounds (e.g., the absence of an active control group). This shortcoming has prevented this approach from being incorporated into educational storytelling (McCabe, 1997). However, Duncker’s paradigm may offer a tool to influence taste preferences and food choices in the especially sensitive kindergarten age. If the paradigm can be shown to work for real food in a setting that appropriately controls possible confounds, the storytelling design could be systematically expanded and accounted for within theoretical frameworks about storytelling.

Therefore, to advance our understanding of narrative effects on food choices, the present study pursued three main aims. Firstly, we wanted to investigate whether the employment of a food-related fairytale-like story can change the food choices of preschool children. Our second aim was to assess the effects of narratives on different types of healthy food choices, that is, fruits and vegetables. Finally, we were interested in the longevity of the intervention effects.

To investigate these questions, we conducted two experiments (Fig. 1), where ad libitum food choices of children aged 4-6 from a platter containing healthy and unhealthy options were tested. In Experiment 1, the healthy food consisted of pieces of fruits (e.g., oranges, bananas), and the non-healthy food consisted of pieces of cakes or cookies. To compare the generalizability of the effects of our stories, in Experiment 2, which was preregistered, the healthy food options were changed to edible raw vegetables, such as pieces of bell pepper or carrots, while the non-healthy food was similar to Study 1. In both studies, after a one-week pre-story baseline measurement, stories were presented, followed by daily measurements of food choices over a two-week (Experiment 1) or three-week (Experiment 2) post-intervention period. To ensure that the demand characteristics were controlled properly, both studies had an active control group that also heard an engaging story structurally similar to our intervention story but food-unrelated. Both narratives contained minimally counterintuitive elements: protagonists in the stories helped the magical painter to refresh the subdued colors of their city either by bringing him healthy food so he gains his strength (in the food-related story) or by providing him with better color (in the food-unrelated story).

**Figure 1.**
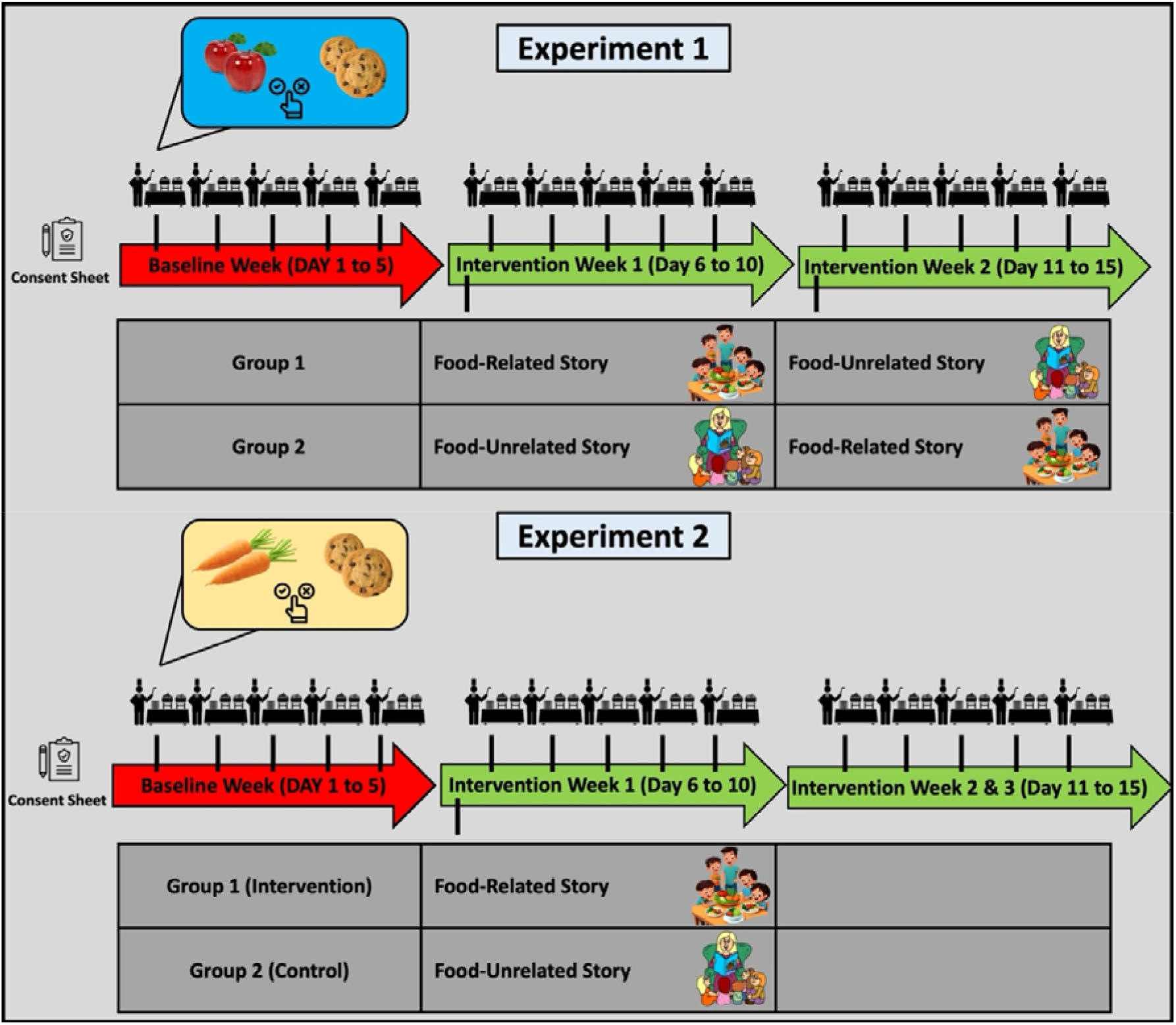
Schematic representation of the experimental procedures. As the experimental manipulation, two interactive stories were used in both studies, one about how two protagonists of similar age as the participants (aged between 4 and 6 years old) saved their hometown from being drained of color with the help of magic fruits or vegetables, while the second control story consisted of a similar plot that did not mention food. Both studies contained one week of baseline measurement, followed by experimental manipulation[s]. In Experiment 1, after the baseline measurement, a crossover design was used, where two groups (*N_total_* = 80) received the food-related story in Week 2 and the food-unrelated story in Week 3 or vice versa. In preregistered Experiment 2, we used a pretest-posttest control group design (*N_total_* = 71) in order to investigate the longevity of the effects. Notably, both food-related and food-unrelated fairytale-like narratives were only presented once on the first day of the targeted week instead of being repeated every day. Participants’ food choices were measured daily by asking them to choose one option from a platter that contained at least two healthy and two unhealthy options. The unhealthy options were always sweet snacks such as cookies. In contrast, although in Experiment 1, healthy options were composed of fruits, in Experiment 2, the healthy options were changed to edible raw vegetables, which are known to be less appetizing than fruits (e.g., Appleton et al., 2016).

## 2. Experiment 1

### 2.1. Methods

#### 2.1.1. Participants

Participants were Kenyan preschool children aged 4 to 6 years (N= 80), drawn from an urban middle-class residential area in Mombasa town. In the second week of the study, participants were assigned to one of two treatment groups of 40 children each, matched for age and the proportion of boys versus girls. The sample size of Experiment 1 was based on convenience, that is, the number of children in the targeted preschool.

The study procedures are in accordance with the Helsinki Declaration and were approved by the ethics committee of the University of Nairobi. Parental consent was obtained for all participants; further, informal assent was obtained from children by giving them the option to either participate or join a play group.

#### 2.1.2. Materials and Tasks

Two stories were developed adhering to principles known to captivate and engross young minds effectively (see appendix). In both stories, on the first day of school, a brother and a sister observe that colors have drained from their city, which is famous for its sparkling colors, and manage to save their city from this disaster. In the food story, the painter who refreshes the colors of the city every night had fallen sick because of eating junk food and was unable to do his job. The children provide him with magic healthy vegetables, after which he regains strength and is able to resume his work. In the control story, the painter used low-quality paint, which could not refresh the sparkling colors of the city; the siblings provided good-quality paint to the painter, and he could restore the colors of the city. During the narration of the stories, there were interactive elements where children talked about junk food and healthy food or about paint and colors. The narration of both stories involved engaging the participants in tasks related to the stories as they were being told (see procedure). Note that no reference to food was made in the control story, which was otherwise, in structure, duration, and interactive elements, very similar to the experimental story.

To test the preference for healthy food over non-healthy food, participants were provided with both healthy and non-healthy food items placed on the same tray. For instance, banana and orange slices would be placed together with cake slices and biscuits, and the children were asked to pick just one item, which they could then eat. Healthy food items consisted of fruits (e.g., apples, bananas, oranges), while the non-healthy food items were pieces of cakes or cookies. There were always at least two healthy food items and two non-healthy ones, but the specific items within each category changed from day to day to maintain variety. In Experiment 1, the food choices were recorded for subgroups of 10 children, that is, for eight subgroups in total. Food items were provided just before the children went for lunch, which is provided in school for all children.

#### 2.1.3. Procedure

During the baseline week, food choices were measured daily in the morning, just before lunch. Afterward, on the first day of the second week, before the food choice test, participants were told one of the stories, depending on their treatment group. Food choice testing commenced for the rest of Week 2 after hearing the story. At the beginning of Week 3 (Day 11), the other story was narrated; that is, children who had been told the food story in Week 2 were now told the paint story and vice versa. Food choice testing then continued until the end of week three.

To ensure effective narration, the kindergarten teachers were engaged in telling the stories, with the goal of captivating their attention while fostering absorption of the stories’ overarching themes.

#### 2.1.4. Data Analysis

By conducting Bayesian generalized linear modeling, we investigated whether stories affected the participants’ food choices (Eq. 1). In the model, choices were predicted based on the Treatment (Baseline, Control, Experimental), the Order of the Treatment (Baseline-Control-Experimental (BCE) vs. Baseline-Experimental-Control (BEC)), and the day of measurement (Day 1 to 5). Based on recent developments in statistics (Rouder et al., 2022; van den Bergh et al., 2023), we used the maximal random effect model. Hence, the full model contained an intercept for the Group and random slopes for Day and Order. As the choices were measured at the sub-group level rather than the individual level, a negative binomial model with log as the link function was used, with the following priors: *N*(0,2.5) as uninformative priors for β coefficients, student_t (3, 0, 2.5) for standard deviations, and student_t (3, 1.6, 2.5) for the intercept of the model. As the full model was multilevel, uninformative priors were preferred (Bürkner, 2017).

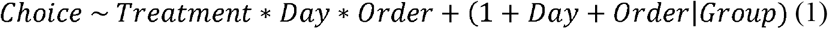

The Bayes factors for the main and interaction effects were calculated similarly to the type-II sum of squares method (van den Bergh et al., 2023) commonly used for the analysis of variance (ANOVA). In other words, for calculating the main effects, a model that contained all main effects and random structure (*Choice~T*+ *D* +*O* + *RE*) was compared to models that did not have the specific main effect but otherwise were the same. Further, the two-way interactions were calculated by comparing a model that consisted of all main effects and two-way interactions (*Choice~T* + *D* + *O* + *T*:*D* + *D*:*O* + *T*:*O* + *RE*) with models that did not contain the specific interaction but otherwise were unaltered. Finally, the three-way interaction was based on a comparison of the full model with a model that did not contain the three-way interaction. Since the inclusion of the interactions depended on the inclusion of the relevant main effects, poorly specified models could not inflate type-I errors (Rouder et al., 2022).

All statistical analyses were conducted using the R programming language (http://www.R-project.org/). For calculating Bayesian hierarchical generalized linear models, brms (Bürkner, 2017) and RSTan (https://mc-stan.org/) were employed. The robust Bayesian correlations were calculated using RStan (https://mc-stan.org/). All models were calculated with five chains, each having 6000 iterations with 2000 warmups. If any variable showed a *Rhat* (i.e., the potential scale reduction factor on split chains) above 1.05, the model was recalculated with increased iterations and reported accordingly. For the model comparison, we used the Pareto smoothed importance sampling (PSIS) estimation of leave-one-out cross-validation (LOO) implemented in the loo package (Magnusson et al., 2020; Vehtari et al., 2016). LOO assesses pointwise out-of-sample prediction accuracy from a fitted Bayesian model using the log-likelihood evaluated at the posterior simulations of the parameter values; however, as it is difficult to calculate, commonly importance weights will be used instead, which results in PSIS-loo. To make sure that PSIS-loo estimation is accurate, one can use *Pareto* 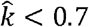 However, *Pareto* 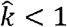 can still be fully trusted.

All hypotheses were tested using the hypothesis package from brms (Bürkner, 2017). Based on the suggestion of van Doorn et al. (2021), *Bayes factors*(*BF*) >3 were considered as significant evidence for the tested hypothesis. One-sided hypotheses (*BF*_+0_ and *BF*_0+_) were the comparison of the posterior probability of hypotheses against their alternative. Further, two-sided tests (*BF*_l0_ and *BF*_0l_) were based on the comparison between hypotheses and their alternative computed via the Savage-Dickey density ratio method (Bürkner, 2017).

### 2.2. Results

By conducting Bayesian generalized linear modeling, we investigated whether stories affected the children’s food choices (Fig. 2). The model converged correctly without any divergent transitions. Further, all *Rhat* = 1, and bulk and tail effective sample sizes were above 9000, showing that model predictions were reliable. Further, the model captured the observed data distribution adequately. The distributions of the beta coefficients can be seen in Figure 1-B.

**Figure 2.**
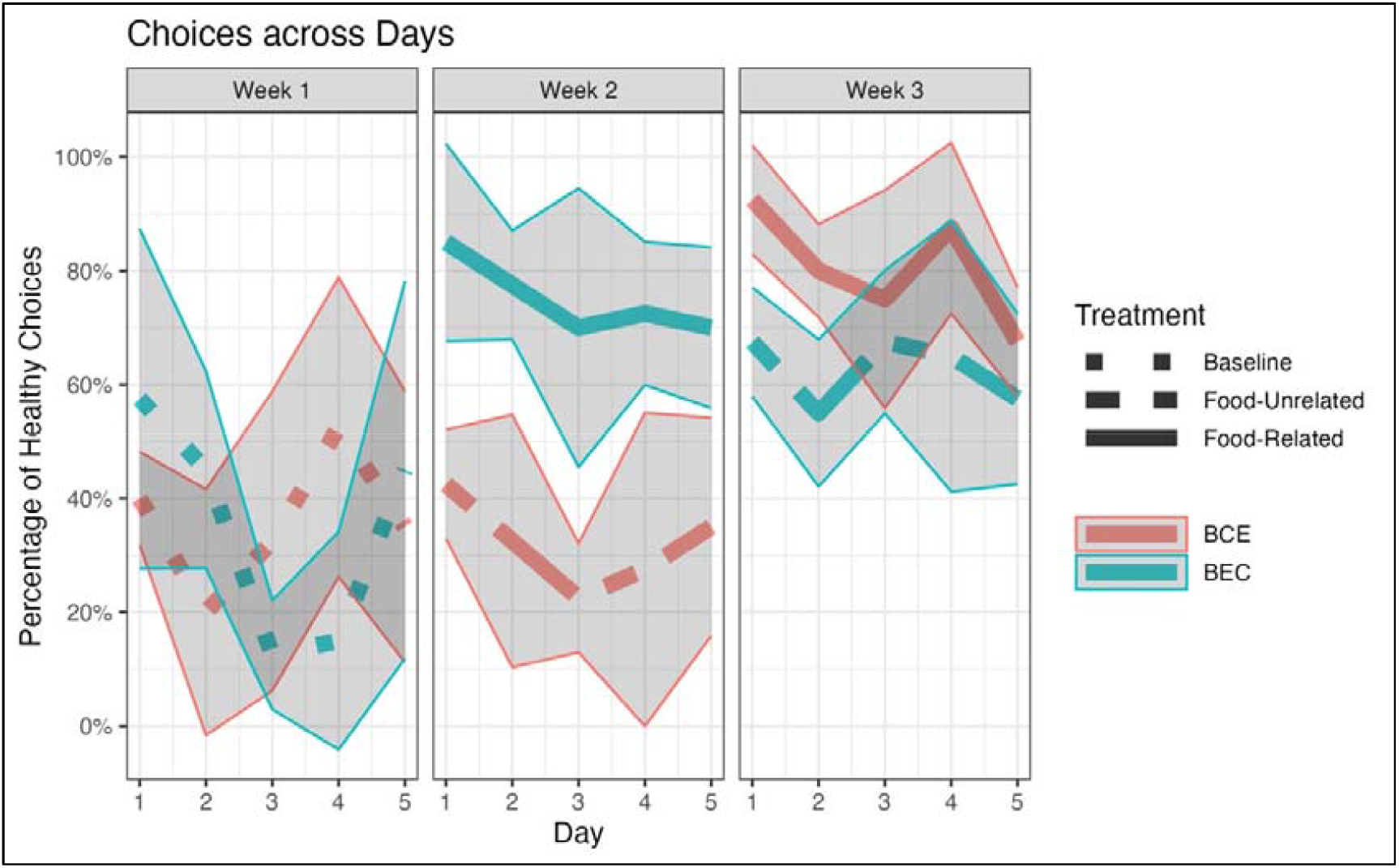
Healthy food choices across days in Experiment 1. The red and blue lines represent the percentage of healthy choices within the two groups with different orders of treatment. Shaded areas represent the standard deviations (SD) of the measured variable. The dotted, dashed, and solid lines represent baseline, food-unrelated narrative, and food-related narrative Weeks, respectively. Narratives were presented on Day 1 of Week 2 and 3, immediately before the food choice assessment.

First, we checked the main effects and interactions via model comparison that resembles the type-II sum of squares method commonly used for ANOVA (Table 1). The effects clearly show that Treatment significantly affected children’s choices (*BF*_l0_ > 9,999). sFollowing up on this result, hypothesis testing revealed that children’s healthy choices ( *mean_Experimental_* = 7.77 ± 1.52; *mean_Control_* = 4.72 ± 2.25; *mean*_*Baseline*_ = 3.55 ± 2.50) were increased after the experimental story compared to the baseline ( H_+_: Treatment_*Experimental*_ > 0; *mean* = 1.12[0.56,1.69], sd = 0.34, *p. p*. > .999, *BF*_+0_ = 2856.1) and the control story ( *H*_+_: Treatment_*Experimental*_ − Treatment_*Control*_ >0; *mean* = 0.9[0.33,1.47], *sd* = 0.25, *p. p*. > .999, *BF*_+0_ = 249). However, the control story did not change children’s choices compared to the baseline (H_+_: Treatment_*Control*_ > 0; *mean*= 0.22[−0.43,0.88], *sd* = 0.4, p. p. = .71, *BF*_+0_ = 2.47). Further, the order of treatments significantly predicted children’s healthy choices (*BF*_l0_ = 181.47), showing that when the experimental (food) story was introduced first, children made more healthy choices during the experiment (*mean*_*BCE*_ = 4.95 ± 2.88; *mean*_*BEC*_ = 5.75 ± 2.61; *H*_+_: Order_BEC_ > 0; *mean*= 0.57[−0.03,1.2], *sd* = 0.39, *p. p*. = .93, *BF*_+0_ = 13.26).

**Table 1.** Fit Indices of the Choice Models computed by multilevel Bayesian modeling.

| Effect | $BF_{10}$ | $\widehat{elpd}_{diff}$ | $se(\widehat{elpd}_{diff})$ |
| --- | --- | --- | --- |
| *Day* | <0.00 | 6.1 | 3.5 |
| ***Order*** | **181.47** | **-2.8** | 1.3 |
| ***Treatment*** | **128489087254.50** | **-29.4** | 6.2 |
| *Day:Order* | 0.02 | 0.7 | 0.6 |
| *Day:Treatment* | <0.00 | 2.2 | 0.5 |
| ***Treatment:Order*** | **16.17** | **-6.5** | 3.2 |
| *Day:Treatment:Order* | 0.01 | -0.8 | 2.7 |

*Note:* Significant effects are indicated in bold. All models are compared to the best model (i.e., Model 1).
$\widehat{elpd}_{loo}$ : Expected log pointwise predictive density for a new dataset using the Pareto smoothed importance sampling (PSIS) leave-one-out cross-validation (loo) criterion (Magnusson et al., 2020; Vehtari et al., 2016). The closer to zero, the better the model is.
$\widehat{elpd}_{diff}$ : The difference between $\widehat{elpd}_{loo}$ of the full model and a model without the specific effect. An effect should be considered to have out-of-sample predictive value if its elimination causes more than two standard error decreases in $\widehat{elpd}_{loo}$ (Magnusson et al., 2020; Vehtari et al., 2016).

The interaction between Treatment and Order was significant (BF_l0_ = 16.17). This result revealed that although there was a significant difference between the experimental and control conditions during the second week ( *H*_-_: Order_BEC_ * Treatment_*Experimental*_ < 0; *mean*= −0.65[−1.4,0.12], sd = 0.46, *p. p*. = .92, *BF*_−0_ = 11.03), this effect was absent during the third week ( *H*_0_: Order_BEC_ * Treatment_*Control*_ =0; *mean*= −0.03[−1.04,0.97], *sd* = 0.51, *p. p*. = .83, *BF*_0l_ = 4.8). These results indicate the longevity of the effects of the experimental story in the absence of a refresher over two weeks. No further interaction or main effect was significant (Table 1).

### 2.3. Discussion

Food choices in Experiment 1 were strongly affected by an interactive, minimally counterintuitive narrative about the benefits of healthy over non-healthy food. After a baseline week of a 30-40% preference for non-healthy items, participants’ choices changed strongly after the food story, where children, on average, chose 80-90% fruits over non-healthy food, which remained stable during the week after the intervention. In contrast, after the control story, no changes in food choice were observed. In Week 3, the food choice of the BEC group was still higher than in the baseline and remained around 60% in favor of fruits. As a demonstration, in Week 3, the food choice in the BCE group, which had remained at baseline during Week 2, increased to ca. 80% preference for healthy choices. These results demonstrate that the effects of fairytale-like narratives persist for at least one full week.

The insensitivity of the groups to the control story (in Week 2 for the BCE group and in Week 3 for the BEC group) is critical as, at these time points, the group hearing the control story served as active controls undergoing the same procedure as the treatment group except for the food-related story content, the presumably active ingredient of the intervention. The replication of the food story effect in the BEC group (Week 3), which was seen in the BEC group (Week 2), suggests that the effect is dissociable from specific participants or experimenter selections. Therefore, the current results strongly imply that the effect of the food story went over and beyond some unspecific confounds, such as motivation, adaptation, or selection bias.

Nevertheless, Experiment 1 leaves open several questions. Would the effects also be present for vegetables, which, in contrast to fruits, are not sweet or harder to chew but are of higher nutritional value? Furthermore, what are the temporal limits of the effect? These questions were addressed in Experiment 2.

## 3. Experiment 2

The preregistered Experiment 2 aimed to replicate and extend Experiment 1. Whereas most elements were the same in both experiments, we used vegetables instead of fruits as healthy food items in Experiment 2. The vegetables offered, such as carrots or bell peppers, were uncooked but commonly considered to be edible in this state. Further, in order to extend the period after the narrative, the experimental or control stories were presented to separate groups (at the beginning of Week 2), and we extended the food choice assessments to three weeks following the narration of the stories. It is also notable that Experiment 2 was conducted in a different kindergarten in a different area of Kenya by a different experimenter, which assesses the generalizability of the results to other samples and procedural variations.

### 3.1. Methods

In reporting the methods, we focus on aspects that differ from Experiment 1.

#### 3.1.2. Participants

The sample size was determined based on the results of Experiment 1 (*N*_*total*_ = 80), where we found a significant main effect of Treatment ( *BF*_l0_ > 9,999), a significant difference between choices after the food story vs. baseline ( *BF*_+0_ = 2856.1) but no difference between the control story vs. baseline (*BF*_+0_ = 2.47). Based on these results, we expected that a similar sample size would result in a 1 − *B* = 99% for the main effect of Treatment. In Experiment 2, we aimed to measure food choices individually rather than in subgroups, which, due to the shrinkage of noise, is expected to result in stronger effects. Therefore, we aimed for the same sample size as in Experiment 1. However, due to the loss of several participants, the final sample size (*N*_*total*_ = 71) was smaller but still satisfied the required power criteria 1 − *B* > 95% (Cohen, 2016). Participants were Kenyan preschool children aged 4 to 6 years and drawn from a middle-class urban area of Machakos county.

After the first week of baseline food choice assessments, participants were assigned to either a control group (N = 35) or an experimental group (N = 36) in Week 2, matched for age and the proportion of boys and girls. The inclusion criteria were similar to Experiment 1.

As in Experiment 1, the study procedures are in accordance with the Helsinki Declaration and were approved by the ethics committee of the University of Nairobi. For all participants, parental consent and their informal assent were obtained prior to the experiment.

#### 3.1.2. Materials and Tasks

Healthy food items consisted of vegetables that are edible in raw form (e.g., bell pepper, carrots, cucumber), while the non-healthy food items were similar to Experiment 1 (i.e., cookies, pieces of cake, etc.). There were always two healthy food items and two non-healthy items, and similar to Experiment 1, the offered items within each category changed from day to day to maintain variety. All other materials and tasks were similar to Experiment 1.

#### 3.1.3. Procedure

The food choices of each participant were recorded individually throughout the study duration. We tested the two groups for four weeks (Week 2 = 5 days; Week 3 = 2 days (Monday, Tuesday); Week 4 = 3 days (Wednesday, Thursday, Friday). Note that some days of food choice assessment had to be skipped due to school holidays. Story narration took place on the first day of Week 2 and was identical in procedure to Experiment 1. Food items were provided just before the children went for lunch, which is provided in school for all children.

#### 3.1.4. Data Analysis

Similar to Experiment 1, by conducting Bayesian generalized linear modeling, we investigated whether the stories differentially affected participants’ food choices (Eq. 2). In the model, choices were predicted based on the Week (Baseline, Manipulation, Longevity), Manipulation type (Experimental vs. Control) and the Day of Measurement (Day 1 to 5). Similar to Experiment 1, we used the maximal random effect model in order to avoid type-I error inflation (Rouder et al., 2022; van den Bergh et al., 2023). Hence, the full model contained an intercept for the participants and random slopes for Day and Week. As the food choices, measured at the individual level, were binary (i.e., 1 for healthy and 0 for non-healthy choices), a Bernoulli model with log as the link function was used, with the following priors: *N*(0,2.5) as uninformative priors for β coefficients, student_t (3, 0, 2.5) for standard deviations, and student_t (3, 0, 2.5) for the intercept of the model. As the full model was multilevel, uninformative priors were preferred (Bürkner, 2017).

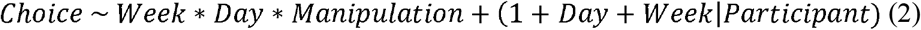

Similar to Experiment 1, Bayes factors for the main effects and interactions were calculated using a method similar to the type-II sum of squares used for ANOVA. In other words, the inclusion of the interactions was dependent on the inclusion of the relevant main effects to prevent poorly specified models from inflating type-I errors.

All statistical analyses were conducted using the R programming language (http://www.R-project.org/). Further, the model calculation and the hypothesis checking were similar to Experiment 1.

### 3.2. Results

To check whether stories affected children’s food choices, we conducted Bayesian hierarchical modeling (Fig. 3). The model converged correctly without any divergent transitions. Further, all *Rhat* = 1, and bulk and tail effective sample sizes were above 5000, showing that model predictions were reliable. Further, the model adequately captured the observed data distribution. The distributions of the beta coefficients can be seen in Figure 2-B.

**Figure 3.**
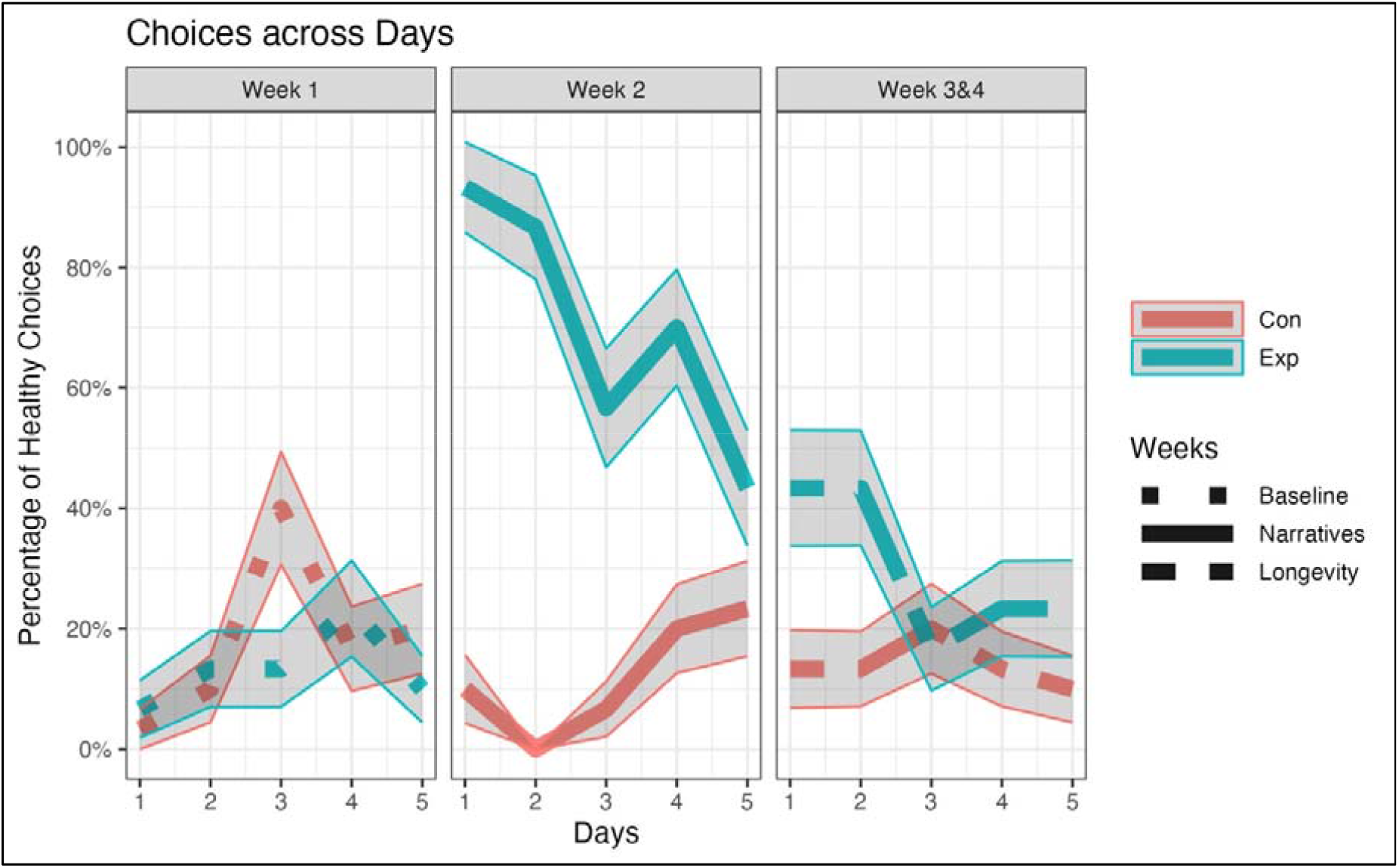
Healthy food choices in Experiment 2 across the 4-week observation period. Red and blue lines represent the percentage of healthy choices within the Control and Experimental groups, respectively. Shaded areas represent the standard deviations (SD) of the measured variable (i.e., sd(*Choices*) = *N_Group_* * *Choice Probability* * [1 − *Choice Probability*]). The dotted, dashed, and solid lines represent baseline, narrative, and longevity Weeks, respectively. Narratives were presented on the first day of Week 2, immediately before the food choice assessment on that day.

First, we checked the main and interaction effects via model comparison resembling the type-II sum of squares method commonly used for ANOVA (Table 2). The effects clearly show that choices were significantly different between Groups (*BF*_l0_ > 9,999), with the experimental group choosing more healthy items (*mean* = 0.32 ± 0.47) compared to the control group (*mean*= 0.12 ± 0.33; *H*_+_: *Group*_*Exp*_ > 0; *mean*= 0.75[−0.44,1.92], *sd* = 0.72, *p. p*. = .85, *BF*_+l_ = 5.7). Further, Week significantly affected participants’ choices ( *BF*_l0_ = 2915; *Week*_*one*_ : *mean* = 0.13 ± 0.34, *Week*_*two*_ : *mean*= 0.35 ± 0.47, *Week*_*three*_ : *mean* = 0.18 ± 0.39). However, there was not enough evidence to reject or accept the difference between either Week One and Week Two (*H*_0_: *Week*_2_ = 0; *mean*= −0.54[−2.16,1.06], *sd* = 0.82, *p. p*. = .72, *BF*_0l_ = 2.61), Week Two and Week Three ( *H*_0_: *Week*_3_ = 0; *mean* = 0.95[−0.49,2.41], *sd* = 2.41, *p. p*.= .6, *BF*_0l_ = 1.52), or Week One and Week Three (H_0_: *Week*_3_ − *Week*_2_ = 0; *mean*= 1.49[−0.2,3.2], *sd* = 0.86, *p. p*. =.47, *BF*_0l_ = 0.9).

**Table 2.**
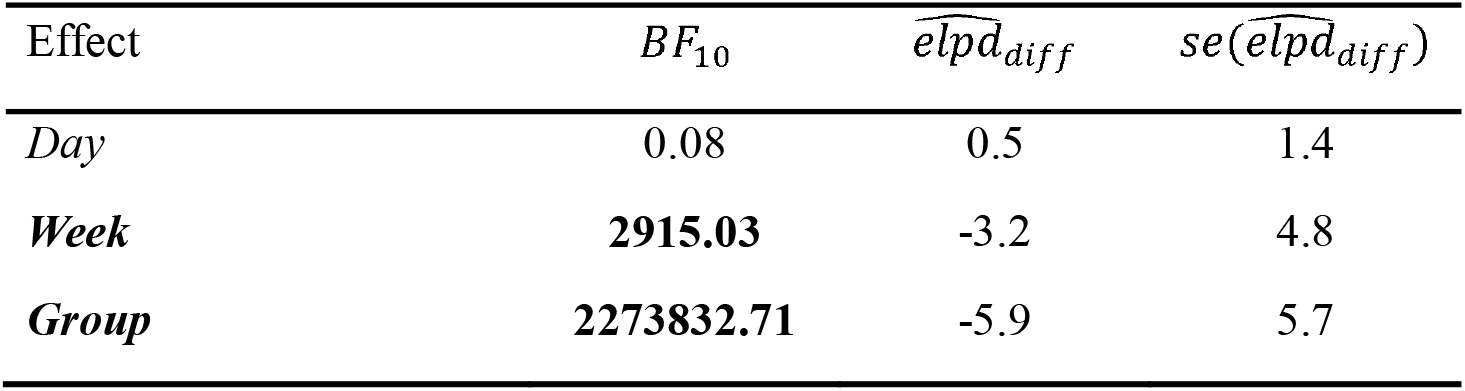
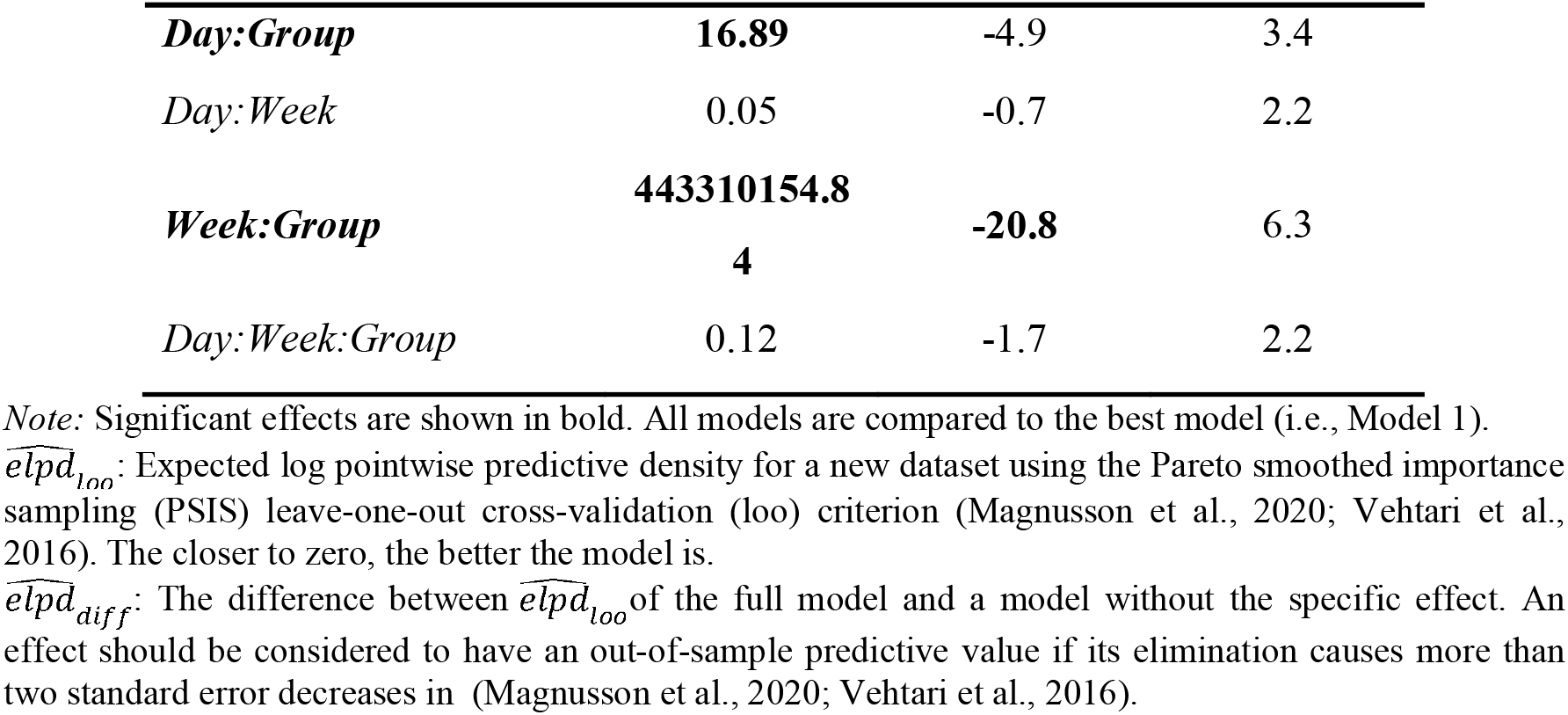
Fit Indices of the Choice Models computed by multilevel Bayesian modeling.

Moving to the interactions, firstly and notably, the interaction between Week and Group was significant ( *BF*_l0_ > 9,999), showing that the experimental compared to the control story positively affected participants’ healthy choices. Following up on this result, the hypothesis testing showed that the increase in healthy food choices in Week 2 compared to Week 1 was significantly higher in the experimental vs. control group ( *H*_+_: *Week*_2_ * *Group*__*Exp*__ > 0; *mean* = 4.59[2.95,6.24], *sd* = 1, *p. p*. > .999, *BF*_+l_ > 9,999). Interestingly, the difference in healthy choices between the two groups was still significant in Week 3 compared to Week 1 ( *H*_+_: *Week*_3_ * *Group*_*Exp*_ >0; *mean*= 0.99[−0.51,2.53], *sd* = 0.92, *p. p*. = .86, *BF*_+l_ = 6.09), indicating that the effect lasts for two weeks.

Further, the interaction between Day and Group was significant ( *BF*_l0_ = 16.8), showing a significant daily decrease in healthy food choices of the experimental group compared to the control group (*H*_-_: Day * *Group*_*Exp*_ < 0; *mean*= −0.28[−0.62,0.07], *sd* = 0.21, *p. p*. = .91, *BF*_+l_ = 10.01). This result indicates that the effect of the experimental story decreased over time. No further interaction or main effect was significant (Table 2).

### 3.3. Discussion

In Experiment 2, we replicated the strong effect of the food story on the food choices of the children but extended it to vegetables. After the same food story as in Experiment 1, the choices of vegetables strongly increased relative to the baseline week and relative to the control group who heard the control story. This is noticeable since it is common to eat uncooked fruits but not so much vegetables (Appleton et al., 2016), and parents often complain that their children do not like eating vegetables. The relative unpopularity of eating raw vegetables is also reflected in the low number of choices for vegetables relative to non-healthy food (cookies, cakes), i.e., participants choose healthy options in approximately 10% of the cases. Therefore, it is all the more impressive that immediately after hearing the food story, the choices of vegetables increased from 10% to 80% and remained above or around 40% for the following eight assessment days. In Week 4 (14-16 days after the intervention), the food choices in the experimental group (i.e., hearing the food story) had returned to baseline. Hence, Experiment 2 indicates the generalizability of the effects of the food story from fruit consumption, observed in Experiment 1, to vegetables and demonstrates the relative longevity of this effect across about two weeks.

## 4. General Discussion

This study had three overarching aims. First, we tested whether a story with counterintuitive elements would influence kindergarten children’s food choices. To this end, we created a food-related fairytale-like narrative focusing on the healthiness and tastiness of fruits and vegetables, which was compared to a control food-unrelated story with a similar structure. Second, we assessed the effectiveness of the food story on two kinds of healthy food, fruits and vegetables, famously known to differ in their appeal to children (Appleton et al., 2016). Third, we investigated the longevity of the effects. Finally, in contrast to previous studies testing the effects of stories on taste preferences (e.g., Duncker, 1938), both experiments benefitted from an active control group that went through the same procedure as the experimental group, except for the presumably active element of the intervention (i.e., a food versus non-food theme). The active control group ensures that the observed results are a specific effect of the food-related content of the story rather than other confounds, such as experimenter effects and arousal.

The most important finding of the present study is that food-related fairytale-like narratives increase choices of both fruits (in Experiment 1) and vegetables (in Experiment 2) compared to baseline and an active control group. The beneficial impact of stories on vocabulary learning and language competence of early readers and preschool children is widely recognized (e.g., Elley, 1989; Ganea et al., 2008; Isbell et al., 2004). These effects seem to be enhanced when combined with guided play (Weisberg et al., 2013), during which adults scaffold child-initiated learning. For pre-primary children, storytelling has been shown to facilitate faster learning and retention through relating and identifying with the characters in stories and developing new perspectives about possibilities in the real world based on story elements (Muindi, 2017). However, as far as food preferences or food choice in children is concerned, there seem to be no published studies on the effects of stories or narratives after the pioneer study of Duncker (1938), which showed an impressive change in liking for unfamiliar tastes after a simple but child-appropriate story with a play-element compared to a no-intervention control group. The current study, however, goes beyond Duncker (1938) by using real food and implementing active control conditions by using a crossover design in Experiment 1 and a random control design in Experiment 2. Although novel, these results are in line with multiple findings in the literature. For instance, it has been shown that the food preferences and choices of young children can be influenced by food advertisements (Emond et al., 2019). Further, in adults, food preferences and choices have been shown to be malleable by direct verbal suggestions (Zahedi et al., 2020, 2023a).

The success of our interactive story intervention on pre-schoolers food choice may relate to its fantastic, minimal counterintuitive elements, such as the idea that colors are restored each night by a painter, that colors fade overnight when the painter is incapacitated, or that magic vegetables and fruits can restore the health of a sick person. From a (controversial) psychoanalytical standpoint, Bettelheim has advocated the importance of fairytales for children (e.g., 2010; for a rebuttal, see Dundes, 1991). The impact of fairytales on children in education from an empirical vantage point has been shown by Weisberg et al. (2015), who found that preschool children learned new words better when they were introduced in the context of a fairytale-like narrative rather than in a more realistic story. According to Muindi (2017), fantasy stories, unlike real stories, foster a state of openness and receptiveness that promotes unconscious learning among pre-primary school learners. While learning is possible through fairytale-like narratives, Hopkins and Weisberg (2017) posit that the story’s success depends on the amount of fantasy and the type of information it conveys. As suggested by Boyer (1984) and demonstrated by Norenzayan et al. (2006), stories containing a minimal violation of a core concept are more memorable than completely intuitive or maximally counterintuitive narratives. In recent neurocognitive studies, Aristei et al. (2022) and Knoop et al. (2024) have shown that minimally counterintuitive concepts interact with emotional content and narrative style. Therefore, it is plausible that the present findings reflect the effects of fantastic stories on children. A further factor contributing to the success of our intervention was the interactive element involved, where children discussed junk food and healthy food. It should also be noted that in the current study, all narrators or proctors were thoroughly familiar to the children because they were their teachers. Therefore, although our control groups have shown that the observed effects cannot be reduced to global demand characteristics imposed by the experimental situations, the familiarity of children with the narrators might have mediated or moderated the observed results (Fairchild & McDaniel, 2017). Therefore, although the current results indicate the effectiveness of the procedure, future research is needed to investigate and verify the effectiveness of different aspects of our intervention.

Additionally, two points need to be discussed here. First, as the narratives used in the current study are minimally counterintuitive, one might ask whether the effects are related to the propensity to form superstitions (Morse, Skinner, 1957) or other supernatural beliefs. It should be considered that the effects observed here are related to a manipulation that does not include any reinforcement. Therefore, these effects should be more attributable to non-reinforced learning (Schonberg & Katz, 2020; Zahedi et al., 2023b), which makes them distinguishable from superstitions that are usually formed based on reinforced learning. Second, as the narratives used in the current study are culturally neutral, that is, a painter refreshes the colors in the city every night to make them brilliant, the observed effects should be minimally counterintuitive across different cultures and thus generalizable to other cultures as well. For instance, Experiments 1 and 2 are conducted in two different regions of Kenya (i.e., Nairobi and Mombasa), which have different religious backgrounds (Christianity-dominated versus Islam-dominated subcultures). Our results show that the effects of our food-related fairytale-like narrative are generalizable across them. However, as the social factors cannot be neglected when considering the effects of direct verbal suggestions (Woody et al., 2005; Zahedi et al., 2024), it is interesting to investigate whether there are any cultural differences beyond normal individual differences observed in response to direct verbal suggestions (Zahedi et al., 2022) in responsiveness to our experimental manipulations.

As our second goal, the two experiments tested the effects of the stories on different types of healthy food, that is, fruits and vegetables. As to be expected, the children had a much lower preference for vegetables than for fruits during the baseline, a finding that has also been shown in other studies (e.g., Appleton et al., 2016). However, interestingly, the story intervention boosted the preference for both types of food similarly. This result is very encouraging since vegetable consumption is considered to be of especially high health benefit (Springman et al. 2020).

The effects of the story were surprisingly long-lasting, considering that the intervention was short (ca. 20 min) and took place only once. In both Experiment 1 and 2, the increase in healthy food choices after the food-related fairytale-like narrative lasted for at least one week. After the intervention, the choice of healthy food soared and stayed elevated for up to two weeks. Notably, the effects of other forms of direct verbal suggestions, such as posthypnotic suggestions, have been shown to be persistent over several weeks in adults (Bohmer & Schmidt, 2022; Zahedi et al., 2023a). Therefore, considering that dietary habits are best shaped at a young age (Scaglioni et al., 2018) and children’s eating behaviors are hard to modify directly (Okely & Hammersley, 2024), the results of our approach are highly promising. Future studies need to address whether the use of multiple or even regular interventions with different stories can extend the effects to a longer period after intervention.

The design of the present study ensured that the effects cannot be attributed to some nonspecific factor. In both experiments, we used active controls that differed from the experimental condition or experimental group only in the presumably active ingredient, which was the association of food (rather than paint) with the rest of the study. Otherwise, we used similar samples in Experiment 2 or even the same sample in Experiment 1, similar environments, the same or similar proctors, and also the stories had the same structure. Hence, one may be confident that the effects of the intervention were isolated from confounds.

During the selection of both healthy and unhealthy items for Experiments 1 and 2, we considered the accessibility and familiarity of food items for children to avoid any food novelty effects on food choices (e.g., Martins & Pliner, 2005; Torri et al., 2020). Additionally, the crossover design for Experiment 1 and the pretest-posttest control group design for Experiment 2 ensured that in both studies, the effects of the experimental group were compared to an active control group. This point further ensures that the observed effects are not related to the selection of items.

Our study is not without limitations. First, we focused mainly on factual food choices rather than food preferences. The rationale for this decision was, however, influenced by the fact that in both experiments, we tested children between 4 and 6 years old, which made conducting interviews or filling out questionnaires rather difficult. Second, we tested our participants only with a limited choice of snacks in small quantities and not real meals, which might be an interesting extension of the approach. Third, in Experiment 1, food choice was assessed in subgroups rather than individually. By using Bayesian generalized linear modeling, we reduced the impact of this limitation in Experiment 1. However, more importantly, in Experiment 2, which was preregistered, we showed that the effects observed in Experiment 1 could be replicated when the choices were measured individually as well. Finally, we measured food choices only once a day via the ad libitum platter; therefore, it might be suggested that the current findings might represent the viability of fairytale-like narratives in experimental conditions rather than real-life spontaneous food consumption. Therefore, these results must be replicated via continuous food consumption measurements to justify real-life applicability.

Future studies should address the generalizability of the current results by focusing on other age groups and more varied food items and presenting the stories in different media such as print, visual, and animations. Additionally, the effects of familiarity of children with narrators should be investigated. Further, future studies should look into the interaction between family settings and the observed effects of stories on children’s food preferences, as not only family setting might be crucial for the long-term success of school-based interventions (Okely & Hammersley, 2024) but also since children have been shown to greatly influence the food environment at their homes (Wingert et al., 2014).

### 4.1. Conclusion

Like other behaviors, food choices are malleable, especially during early childhood. The current study showed that only hearing a fairytale-like narrative about healthy food on a single occasion can modulate food choices for at least two weeks. In general, the effects generalized from fruits to vegetables were replicable over two samples and surprisingly stable, considering the relatively short experimental manipulation that was used (only 20 minutes). The story effects may be partly attributed to their minimally counterintuitive elements (Weisberg et al., 2015), as it has been shown that fairytale-like narratives are processed more deeply compared to stories that fully comply with core knowledge about the world (Aristei et al., 2022; Knoop et al., 2024). In summary, these results point to the powerful effects of fairytale-like narratives to alter food preferences in early childhood at a time when unhealthy eating is becoming a pandemic.

## Data Availability

All data produced are available online at OSF.

https://doi.org/10.17605/OSF.IO/7AN24

https://doi.org/10.17605/OSF.IO/CMDAH

